# Rapid spread of the SARS-CoV-2 Omicron XDR lineage derived from recombination between XBB and BA.2.86 subvariants circulating in Brazil in late 2023

**DOI:** 10.1101/2024.05.07.24306998

**Authors:** Ighor Arantes, Kimihito Ito, Marcelo Gomes, Felipe Cotrim de Carvalho, Walquiria Aparecida Ferreira de Almeida, Ricardo Khouri, Fabio Miyajima, Gabriel Luz Wallau, Felipe Gomes Naveca, Elisa Cavalcante Pereira, COVID-19 Fiocruz Genomic Surveillance Network, Marilda Mendonça Siqueira, Paola Cristina Resende, Gonzalo Bello

## Abstract

Recombination plays a crucial role in the evolution of SARS-CoV-2. The Omicron XBB* recombinant lineages are a noteworthy example, as they have been the dominant SARS-CoV-2 variant worldwide in the first half of 2023. Since November 2023, a new recombinant lineage between Omicron subvariants XBB and BA.2.86, designated XDR, has been detected mainly in Brazil. In this study, we reconstructed the spatiotemporal dynamics and estimated the absolute and relative transmissibility of the XDR lineage. The XDR lineage displayed a recombination breakpoint in the ORF1a coding region, and the most closely related sequences to the 5’ and 3’ ends of the recombinant correspond to JD.1.1 and JN.1.1 lineages, respectively. The first XDR sequences were detected in November 2023 in the Northeastern Brazilian region, and their prevalence rapidly surged from <1% to 25% by February 2024. The Bayesian phylogeographic analysis supports that the XDR lineage likely emerged in the Northeastern Brazilian region around late October 2023 and rapidly disseminated within and outside Brazilian borders from mid-November onwards. The median effective reproductive number of the XDR lineage in Brazil during the initial expansion phase was estimated to be around 1.5. In contrast, the average relative instantaneous reproduction numbers of XDR and JN* lineages were estimated to be 1.37 and 1.29 higher than that of co-circulating XBB* lineages. In summary, these findings support that the recombinant lineage XDR arose in the Northeastern Brazilian region in October 2023, shortly after the first detection of JN.1 sequences in the country. In Brazil, the XDR lineage exhibited a higher transmissibility level than its parental XBB.* lineages and is spreading at a rate similar to or slightly faster than the JN.1* lineages.

## INTRODUCTION

Recombination is a major driving force of virus evolution and has contributed significantly to the genetic diversity of SARS-CoV-2 lineages (Turakhia et al., 2022). The first documented example of a SARS-CoV-2 variant increasing its fitness through recombination, rather than substitutions, was the Omicron subvariant XBB that emerged through the recombination of two cocirculating BA.2 lineages, BJ.1 and BM.1.1.1, around mid-2022 (Tamura et al., 2023). The XBB lineage showed a significant capacity to evade population immunity and a substantially higher effective reproduction number (R_e_) than the parental lineages, suggesting that the recombination event increased viral fitness (Tamura et al., 2023). In early 2023, multiple XBB descendent lineages, such as XBB.1.5*, XBB.1.9.2*, and XBB.1.16*, spread worldwide and rapidly became predominant (Chen et al., 2021). As a result, in May 2023, the World Health Organization (WHO) Technical Advisory Group on COVID-19 Vaccine Composition recommended using a monovalent XBB.1 descendent lineage, such as XBB.1.5, as the vaccine antigen (WHO, 2023). The upsurge of the XBB lineages highlights the importance of real-time analyses of viral genomes to monitor the potential emergence of novel SARS-CoV-2 recombinant lineages with higher transmissibility, virulence and/or immune scape properties to guide vaccine composition updates.

In August 2023, a highly mutated Omicron subvariant named BA.2.86, with over 30 mutations in the spike (S) protein concerning BA.2 and XBB subvariants, was identified and classified by the WHO as a variant under monitoring (WHO, 2020). This novel subvariant probably evolved through a saltation-like process directly from a BA.2 ancestor circulating in South Africa around May 2023 (Khan et al., 2023). The average R_e_ of BA.2.86 was estimated to be about 1.29-fold greater than that of XBB.1.5 and 1.07 higher than that of EG.5.1, suggesting that BA.2.86 potentially has remarkable fitness considering the XBB variants (Tamura et al., 2024; Uriu et al., 2023). The BA.2.86 subvariant, however, was not as immune evasive as the XBB lineages circulating in mid-2023 and failed to become dominant at a global scale (Khan et al., 2023a; Uriu et al., 2023; Q. Wang et al., 2023; Qu et al., 2024; Zhang et al., 2024; Hu et al., 2023; Lasrado et al., 2023; Planas et al., 2024). Instead, the BA.2.86 sub-lineage JN.1, with just one additional S mutation (S:L455S), rapidly surged on a global scale in late 2023 and was designated by the WHO as a variant of interest (VOI) in December 2023 (Looi, 2023; X. Wang et al., 2024). JN.1 displayed lower affinity to ACE2 but enhanced immune escape compared with its predecessor BA.2.86 (Yang et al., 2024; Kaku et al., 2024b; Planas et al., 2024), which partly explains the higher R_e_ of JN.1 with respect to that of BA.2.86.1 and XBB lineages co-circulating in several European countries at the end of November 2023 (Kaku et al., 2024).

The SARS-CoV-2 epidemic in Brazil in 2023 was driven by the spread of different XBB lineages that locally evolved by the stepwise accumulation of mutations in the S protein, which increased the affinity to the angiotensin-converting enzyme 2 (ACE2) receptor, or the immune escape (Arantes et al., 2024). Genomic data compiled by the COVID-19 Fiocruz Genomic Surveillance Network (https://www.genomahcov.fiocruz.br/dashboard-en/) indicates that XBB lineages dominated the Brazilian epidemic until October 2023 but have been progressively outcompeted by the VOI JN.1.* since November 2023. Interestingly, that data also revealed the concomitant spread of a new Omicron subvariant in Brazil, XDR, that arose through recombination between XBB and BA.2.86 cocirculating subvariants. In this study, we explored the origin, dissemination pattern, and transmissibility of this novel XDR recombinant lineage between November 2023 and February 2024.

## METHODS

### SARS-CoV-2 Brazilian genome sequences

A total of 3,425 SARS-CoV-2 complete genome sequences recovered across 19 Brazilian states between October 1^st^, 2023, and February 29^th^, 2024, were newly generated by the COVID-19 Fiocruz Genomic Surveillance Network (https://www.genomahcov.fiocruz.br/en/). All samples had real-time RT-PCR cycling threshold (Ct) below 30. There was no intended bias considering dense sampling of specific outbreaks, and we do not consider the health status (severe, mild, or asymptomatic) or epidemiological data (gender and age) of individuals as a criterion for sampling. SARS-CoV-2 genome sequences were generated using the COVIDSeq Test (Illumina) and ARTIC 4.1v primers set or 5.3.2v NCOV 2019 panel (IDT), as previously described (Resende et al., 2021). Raw data were converted to FASTQ files at Illumina BaseSpace cloud, and consensus sequences were produced with the DRAGEN COVID Lineage v4 or ViralFlow 1.0 (Dezordi et al., 2022). All genomes were uploaded to the EpiCoV database of GISAID (https://gisaid.org/). Additionally, all SARS-CoV-2 complete genomes (> 29,000 nucleotides) collected in the country since October 1^st^, 2023, and submitted to the EpiCov database until February 29^th^, 2024, with complete collection date, lineage assignment, and limited presence of unidentified positions (N < 5%) were downloaded (n = 2,727). Whole-genome consensus sequences were classified using the ‘Phylogenetic Assignment of Named Global Outbreak Lineages’ (PANGOLIN) software v4.3.1 (pangolin-data version 1.21) (Rambaut et al., 2020; O’Toole et al., 2021).

### XDR dataset and maximum likelihood (ML) analysis

To comprehensively analyze the spatiotemporal dynamics of the SARS-CoV-2 XDR lineage, we compiled a dataset comprising all Brazilian (n = 161) and non-Brazilian (n = 25) sequences attributed to this lineage collected from October 1, 2023, to February 29, which were submitted to the EpiCov database with the previously described parameters. The resulting dataset [n = 186; 29,421 nucleotides from position 1 of ORF1ab to position 117 of ORF10 (Okada et al., 2020)] was aligned using MAFFT v7.467 (Katoh et al., 2017). To identify the XBB and BA.2.86 sub-lineages that serve as parental to XDR, we created a consensus from the complete XDR dataset using Seaview software v.5.0.5 (Gouy et al., 2009) with a 90% identity threshold. From this consensus, the initial 11,043 positions related to XBB and the final 17,695 positions associated with XBB.2.86 were used in two local blastn (Altschul et al., 1990) searches. Reference datasets for these experiments were compiled by downloading all XBB.1.5* (n = 18,303) and JN.1* (n = 1,898) genomes from the EpiCov database, sampled in the two months prior to the first detection of XDR on November 8^th^, 2024. We selected the top 500 hits from each search, aligned them to their corresponding subgenomic XDR sequence, and conducted a maximum likelihood (ML) phylogenetic analysis using IQ-TREE v2.2.2.7 (Minh et al., 2020) under the best nucleotide substitution model (GTR+I+G), as selected by the ModelFinder application (Kalyaanamoorthy et al., 2017) embedded in IQ-TREE. The approximate likelihood-ratio test (aLRT) (Anisimova & Gascuel, 2006) assessed the branch support based on the Shimodaira–Hasegawa-like procedure with 1,000 replicates. An ML analysis was also performed on the entire XDR dataset using the same parameters, and the resulting tree was selected for further analysis.

### Spatiotemporal dynamics of the XDR lineage

The temporal signal of the XDR complete dataset was assessed from the previously inferred ML tree by performing a regression analysis of the root-to-tip divergence against sampling time using TempEst v1.5.36 (Rambaut et al., 2016). Sequences that diverged more than 1.5 interquartile ranges from the root-to-tip regression were considered outliers and removed from subsequent analysis. The significance of the association between the two variables was assessed with a Spearman correlation test implemented in the R programming language v.4.1.2 (R Core Team, 2021), along with its base package. A time-scaled Bayesian phylogeographic tree was estimated in BEAST v1.10.4 (Rambaut, 2000; Suchard et al., 2018) with BEAGLE (Suchard & Rambaut, 2009) to improve run-time, under a relaxed uncorrelated molecular clock model (Drummond et al., 2006) with a uniform distribution between 5.0E-4 and 1.5E-3 (Duchene et al., 2020; Ghafari et al., 2022; Tay et al., 2022), the non-parametric Bayesian Skyline model (BSKL) (Drummond, 2005) as the coalescent tree prior, and a reversible discrete phylogeographic model (Lemey et al., 2009). The Brazilian sequences were categorized based on their regions of origin (n = 5), while foreign ones were grouped according to their subcontinental regions (n = 3). Markov chain Monte Carlo (MCMC) simulations were run sufficiently long to ensure convergence [effective sample size (ESS) > 200] in all parameters as assessed in TRACER v1.7 (Rambaut et al., 2018). Additionally, the number and directionality of location transitions in XDR history were estimated with a Markov Jumps count (O’Brien et al., 2009). The maximum clade credibility trees were summarized with TreeAnnotator v1.10 (Suchard et al., 2018) and visualized using Treeio v3.1.7 (Wang et al., 2019) and ggtree v3.2.1 R packages (Yu, 2020). Graphs used to present the results were generated using the ggplot2 R package (Wickham, 2016).

### Effective reproductive number (R_e_) estimation

The temporal trajectory of the R_e_ of the XDR lineage in Brazil was estimated from genomic data by using the Birth-Death Skyline model (BDSKY) model (Stadler et al., 2012) implemented within BEAST 2 v.2.6.2 (Bouckaert et al., 2019). The sampling rate was set to zero for the period before the oldest sample and estimated from the data afterward. The BDSKY prior settings were as follows: become uninfectious rate (exponential, mean = 36); reproductive number (log-normal, mean = 0.8, s.d. = 0.5); sampling proportion (beta, alpha = 1, beta = 275). The origin parameter was conditioned to root height, and R_e_ was estimated in a piecewise manner over four time intervals defined from the date of the most recent sample up to the root of the tree. A normal prior was applied to the T_MRCA_ of the lineage based on the estimations performed during time-scale analysis. The MCMC chain was run until all relevant parameters reached ESS >200, as previously explained. The inferred temporal evolution of the R_e_ was plotted with the bdskytool R package (https://github.com/laduplessis/bdskytools).

### Relative instantaneous reproduction number (R_RI_) estimations

We measured the transmission advantage of JN.* and XDR lineages compared to the XBB.* lineages cocirculating in Brazil by estimating the R_RI_ from the observed frequencies of variants in the country from November 2023 to January 2024 as described elsewhere (Ito et al., 2021). The probability mass function of the generation time of Omicron was modeled by discretizing the gamma distribution with α=4.03 and θ=0.737, so that the gamma distribution has the same mean (2.97) and variance (2.19) as the log-normal distribution estimated by Park et al. (Park et al., 2023). SARS-CoV-2 lineages observed in each country region were counted using half-month bins. Sequences from Ceara and Bahia were excluded as these states displayed a singular epidemiological molecular pattern within Brazil characterized by very early spread of JN.1* lineages and a high prevalence of the JN.1.23 lineage, respectively (data not shown). By maximizing the likelihood function of the multinomial distribution, we estimated the R_RI_ of JN.* and XDR lineages with respect to (w.r.t.) the XBB.* lineages and initial frequencies of variants on November 1, 2023, using datasets from Brazil, or its Northeastern, Southeastern, and Centra-Western regions. The 95% confidence intervals (CI) of estimates were estimated by profile likelihood. The trajectories of variant frequencies were calculated by using the maximum likelihood estimates of the parameters in the model. The 95% CI of the trajectories was calculated by using combinations of parameters within the 95% confidence region (Held & Sabanés Bové, 2020).

### Data on hospitalizations for severe acute respiratory illness (SARI)

We extracted data about hospitalizations resulting from SARI attributed explicitly to SARS-CoV-2 (SARI-COVID) in Brazil from November 2023 to January 2024. This information was sourced from the Influenza Surveillance Information System (SIVEP-Gripe) database (https://opendatasus.saude.gov.br/dataset?tags=SRAG), as previously described (Arantes et al., 2024b).

## RESULTS AND DISCUSSION

Among SARS-CoV-2 nearly complete genome sequences submitted to the EpiCoV database of GISAID until 5^th^ March 2024, we identified 203 sequences classified as XDR from 8^th^ November 2023 onwards. Most of the XDR sequences were sampled in Brazil (87%), including the earliest sequences sampled. The remaining XDR sequences were sampled in neighboring South American countries (Argentina, Chile, Paraguay, and Uruguay, 6%), North America (Canada and USA, 4%), and Europe (Denmark and Spain, 2%). The XDR recombinant genomes contain all the JD.1.1 and none of the JN.1.1 lineage-defining mutations until the mutation ORF1a:V3593F, and all JN.1.1 lineage-defining mutations starting from the mutation ORF1a:R3821K, thus supporting a recombination breakpoint between nucleotide positions 11,043 and 11,726 of the ORF1a coding region (**Fig. 1a**). In order to confirm the parental lineages of the XDR recombinant, we performed a BLAST search analysis to detect the most similar sequences to the 5’ and the 3’ ends flanking the recombination breakpoint among XBB.1.5* and JN.1* sequences globally sampled over two months before the detection of the first XDR genome and that were submitted to the GISAID database until 5^th^ March 2024. The ML phylogenetic analysis supports that the most closely related sequences to the XDR recombinant’s 5’ and 3’ ends correspond to JD.1.1 and JN.1.1 lineages, respectively (**Fig. S1a-b**).

**Figure 1.**
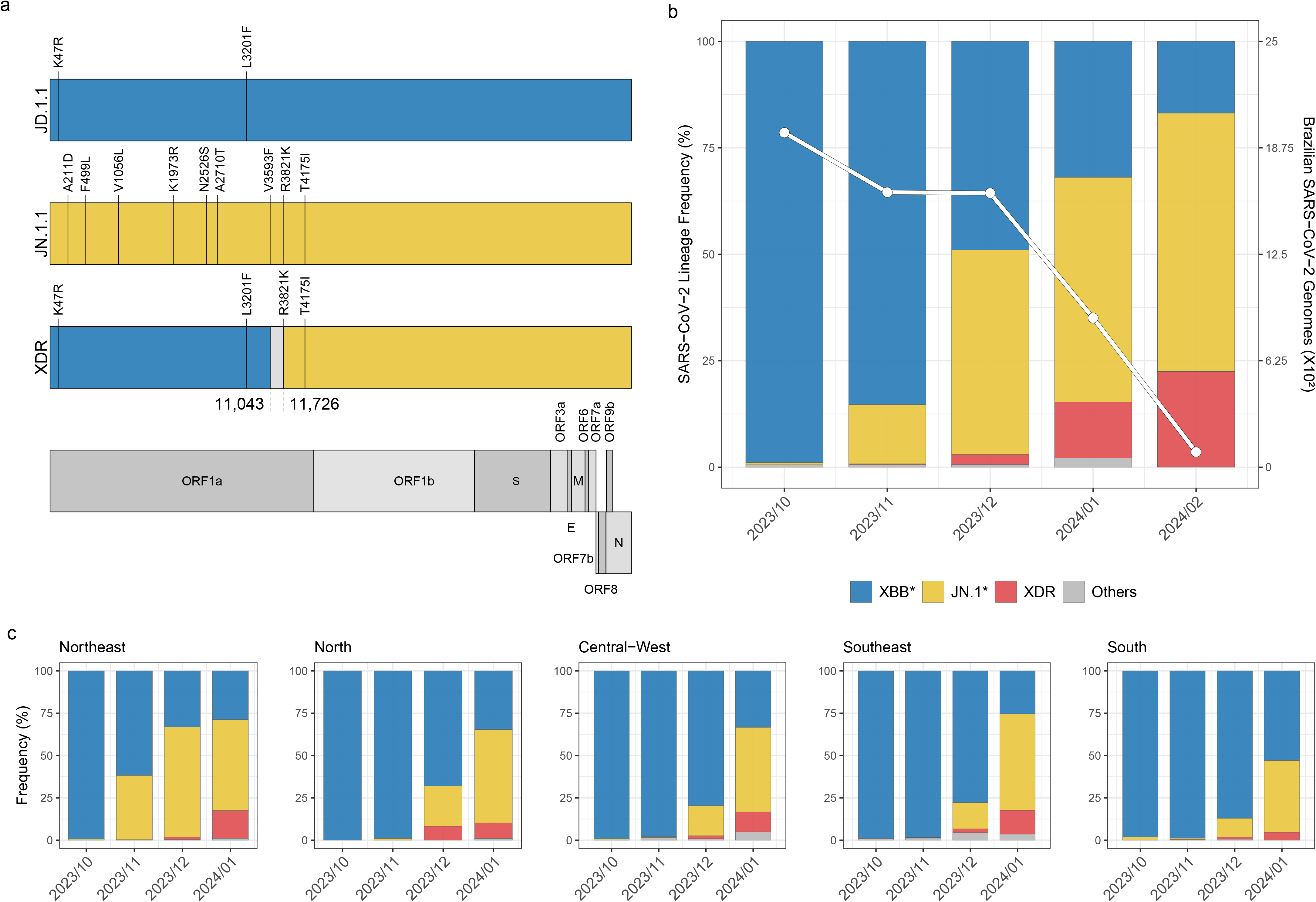
Genomic structure and prevalence of SARS-CoV-2 XDR lineage in Brazil. (**a**) Recombinant structure of the SARS-CoV-2 XDR lineage. Synapomorphic amino acid mutations of the XDR lineage and the most probable parental lineages (JD.1.1 and JN.1.1) compared to the BA.2 lineage are highlighted. The genomic coordinates within ORF1a that flank the recombination point are also annotated. Each segment of the XDR genome is color-coded based on its parental lineage, with the segment that potentially houses the recombination point indicated in light gray. (**b**) Relative prevalence of the XDR lineage in Brazil’s SARS-CoV-2 epidemic between October 2023 and February 2024 is shown alongside the monthly count of viral genomes sampled in the country (n_TOTAL_ = 6,152). (**c**) Relative prevalence of the XDR lineage stratified by the country’s regions using the same color code provided in the previous panel. The main lineage groups considered (XBB*, JN.1.*, and XDR) are colored according to the legend in the previous panel.

In order to describe the changing molecular pattern of the SARS-CoV-2 epidemic in Brazil in recent months, we analyzed 6,152 Brazilian SARS-CoV-2 genomes collected between 1^st^ October 2023 and 29^th^ February 2024 (**Fig. 1b**). As of October 2023, most SARS-CoV-2 sequences in Brazil belong to the XBB lineages (**Fig. 1b**). The first JN.1.* lineages were identified in October 2023 across the Central-Western and Southern regions and their prevalence steadily rose from <1% in that month to 60% in February 2024 (**Fig. 1b**). Concurrently, the first XDR sequences were detected in November 2023 in the Northeastern region and their prevalence surged from <1% in that month to 25% by February 2024, marking a significant increase in the frequency of this recombinant lineage in the study period (**Fig. 1b**). The same overall pattern of substitution of lineages XBB by JN.1.* and XDR was consistently observed in the whole country over last months, although the current prevalence of the different Omicron subvariants vary across Brazilian regions (**Fig. 1c**). The expansion of the JN.1.* and XDR lineages started in the Northeastern region and culminated in the Southern region. By January 2024, the XBB lineages only persisted as the predominant Omicron subvariant in the Southern region, comprising approximately 55% of cases.

We conducted a Bayesian phylogeographic inference to ascertain the spatiotemporal dissemination pattern of the XDR lineage. After removing low-quality sequences with high frequency of private mutations or missing data of the entire receptor-binding domain of the S protein, we retained a total of 186 XDR sequences from Brazilian Northeastern region (41%), Brazilian Southeastern region (9%), Brazilian Central-Western region (22%), Brazilian Northern region (13%), Brazilian Southern region (2%), other South American countries (7%), North America (4%) and Europe (2%). The regression analysis of the root-to-tip divergence against sampling time revealed a satisfactory temporal signal in the XDR dataset (**Fig. 2a**), and the Bayesian time-scaled phylogenetic analysis estimated an evolutionary rate of 7.5 x 10^-4^ (95% HPD: 5.1 -9.7 x 10^-4^) substitutions/site/year, consistent with that inferred for other SARS-Cov-2 lineages (Duchene et al., 2020; Ghafari et al., 2022; Tay et al., 2022). The Bayesian phylogeographic analysis indicates that the XDR lineage likely arose in the Northeastern region (*PSP* = 0.99) around late October 2023 (**Fig. 2b**). From mid-November 2023 onwards, the XDR lineage spread mainly from the Northeastern to other Brazilian regions and beyond Brazil’s borders (**Fig. 2c-d**).

**Figure 2.**
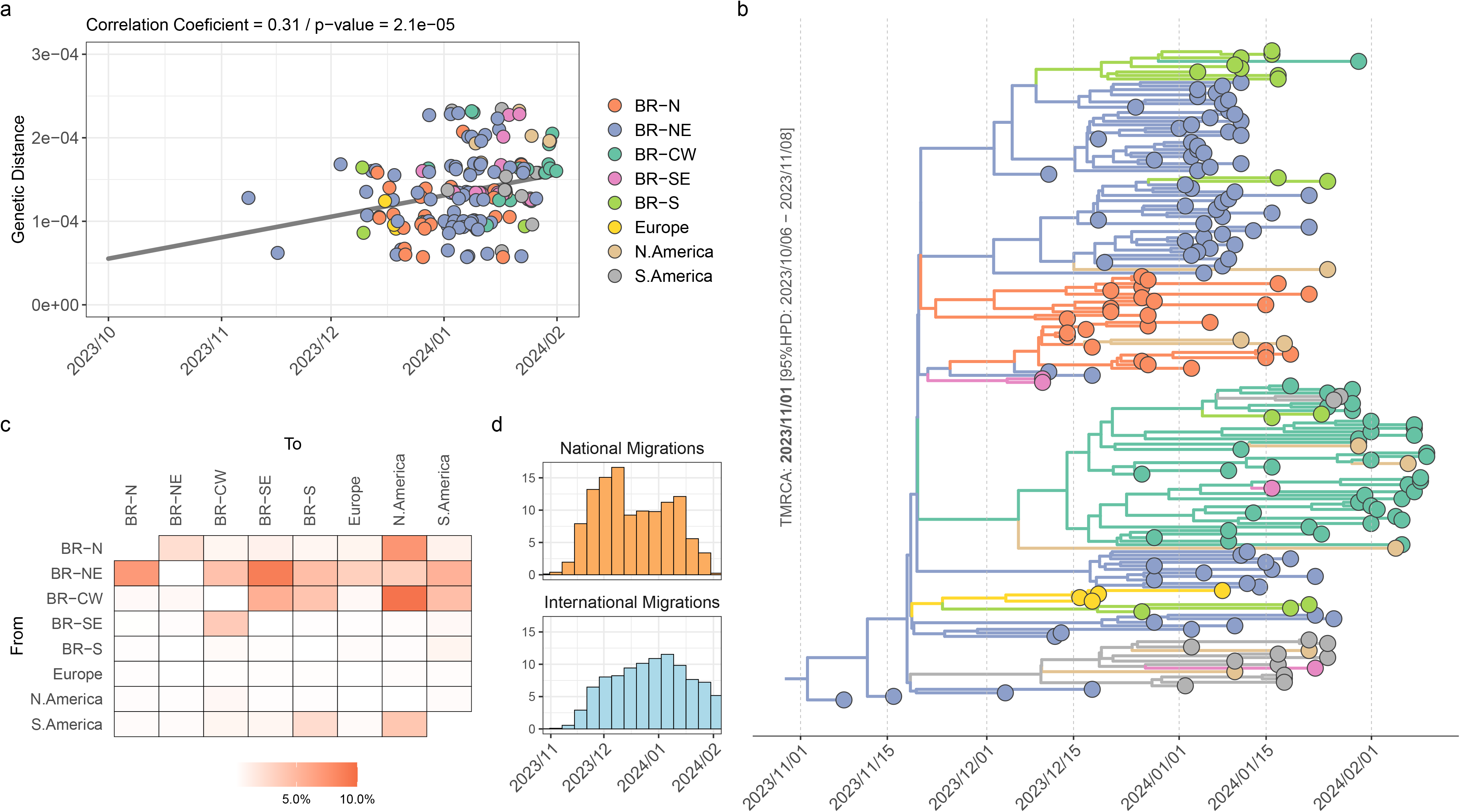
Spatiotemporal dynamics of global dissemination of SARS-CoV-2 XDR lineage. (**a**) Plot of the root-to-tip divergence against collection dates of XDR genomes sampled worldwide (n = 186) between November 2023 and February 2024. The correlation coefficient and p-value obtained in the analysis are annotated at the top of the panel. Data points are colored according to the sampling location, as indicated in the legend. (**b**) Time-scale maximum clade credibility Bayesian tree of the XDR lineage. Tips and branches colors indicate the sampling state and the most probable inferred state of the nodes, respectively, as indicated in the legend. The inferred T_MRCA_ of the lineage is annotated near the root node of the tree. All horizontal branch lengths are time-scaled, and the tree was automatically rooted under the assumption of the molecular clock model. (**c**) Heatmap cells are colored according to the estimated number of viral migrations between locations based on the Markov Jump counts in the Bayesian phylogeographic analysis. (**d**) Estimated number of viral migrations among Brazilian regions (National Migrations) and between any Brazilian region and any foreign country (International Migrations) through time. BR: Brazil, N: North, NE: Northeast, CW: Central-west, SE: Southeast, S: South.

We then applied the BDSKY model to estimate the R_e_ of the XDR lineage by selecting all sequences from Brazil. The temporal trajectory of the median Re supports an initial expansion phase (R_e_ ∼ 1.5) during November and December 2023, followed by a phase of stabilization (R_e_ ∼ 1) in January 2024 and subsequent decrease (R_e_ < 1) in February 2024 (**Fig. 3a, Table S1)**. Interestingly, the trajectory of the R_e_ of the XDR lineage coincides with the overall pattern of severe acute respiratory illness (SARI) cases detected in the Northeastern Brazilian region during that period (**Fig. 3a)**. We speculate that the recent dissemination of the XDR lineage outside the epicenter was probably not captured by our BDSKY model and that more sequences collected at recent times are probably necessary to accurately estimate the R_e_ of the XDR lineage at February 2024. The median R_e_ of the XDR lineage in late 2023 was lower than those estimated for different SARS-CoV-2 lineages spreading in distinct Brazilian regions during epidemic waves in 2020-2022, such as the B.1.* (R_e_ = 2.0–3.8), Gamma P.1 (R_e_ = 2.0-2.6), and Omicron BA.1 (R_e_ = 2.5–3.0) (Resende et al., 2021; Naveca et al., 2021; Arantes et al., 2024). This supports that current Brazilian population immunity probably confers some level of protection against reinfections by XDR (and also probably JN.1*) lineages.

**Figure 3.**
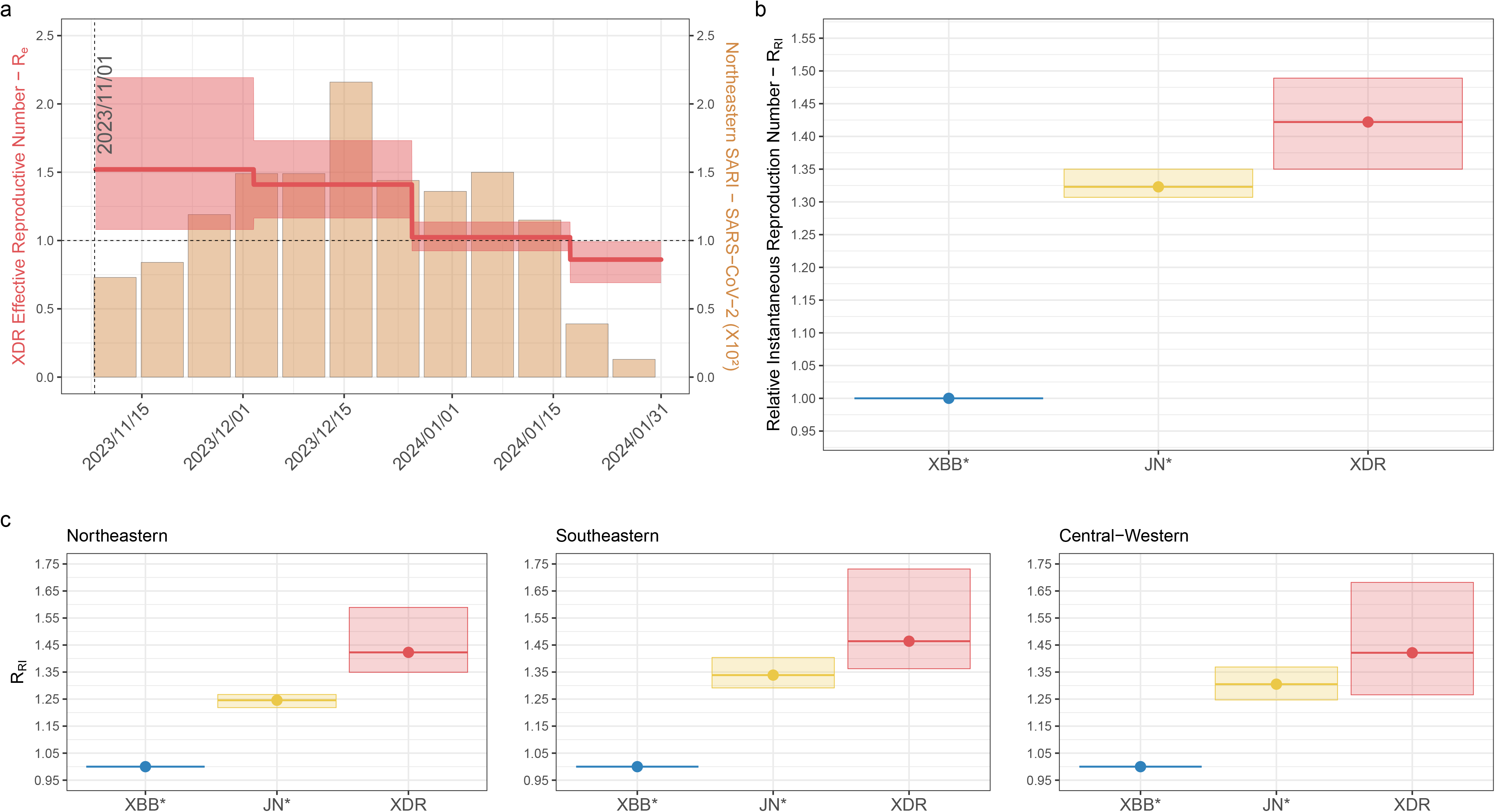
Absolute and relative R_e_ of SARS-CoV-2 XDR lineage in Brazil. (**a**) Temporal variation in the R_e_ of the XDR lineage circulating in Brazil was estimated using the BDSKY approach, juxtaposed with the monthly count of SARI cases attributed to SARS-CoV-2 in the Brazilian Northeastern region. The median (solid red line) and the 95% HPD interval (shaded red area) of the R_e_ estimates are represented together with the estimated T_MRCA_ of the lineage (vertical shaded line). (**b-c**) Relative transmissibility (R_RI_) of XDR and JN.* lineages w.r.t. the XBB.* lineages estimated from the observed frequencies of variants circulating in Brazil (**b)** or in specific country regions **(c**). The average (solid lines) and the 95% CI interval (shaded area) of the R_RI_ estimates are represented.

To trace the shift in viral fitness associated with the emergence of the XDR recombinant, we estimated the R_RI_ of XDR and JN.* (BA.2.86 + S:L455S) lineages w.r.t. XBB.* lineages cocirculating in Brazil from the observed frequencies of variants in the country from November 2023 to January 2024. These analyses support that JN.* lineages were more transmissible than XBB lineages and that the XDR recombinant lineage showed higher transmissibility than their parental lineages. The average R_RI_ of JN.* and XDR lineages in Brazil were estimated to be 1.29- and 1.37-times higher than that of XBB.* cocirculating lineages (mainly JD.1* and GK.1*), respectively (**Fig. 3b)**. Thus, the average R_RI_ of XDR lineage was 1.06-times higher than that of JN.* lineages. We also obtained specific R_RI_ values for the Northeastern, Southeastern, and Central-Western Brazilian regions. All analyses point to a higher transmissibility of the XDR lineage w.r.t. the XBB.* lineages across all country regions and w.r.t. the JN.* lineages only in the Northeastern region (**Fig. 3c)**. Interestingly, the average R_RI_ of JN.* and XDR lineages w.r.t. XBB lineages co-circulating in Brazil in late 2023, were quite similar to the average R_RI_ of XBB.* lineages w.r.t. BA.5 lineages (mainly BQ.1*/BE*) co-circulating in Brazil in early 2023 (R_RI_ = 1.24-1.48) (Arantes et al., 2024), and was also similar to the estimated relative R_e_ of JN.1 w.r.t. XBB lineages co-circulating in Europe such EG.5.1, HK.3 and JD.1* (∼1.10-1.40) (Kaku et al., 2024).

In summary, our analyses suggest that the XDR lineage likely originated from a recombination event between SARS-CoV-2 lineages JD.1.1 and JM.1.1 in the Northeastern region of Brazil in late October 2023 and was detected just a couple of weeks after its emergence. The XDR lineage spread to other Brazilian regions and beyond Brazil’s borders from the Northeastern region. Our findings suggest that the XDR recombinant lineage exhibited higher transmissibility than both parental lineages, particularly in the Northeastern region. This result is intriguing as both the XDR and JN.1* lineages have identical S protein sequences, raising the possibility of viral transmissibility factors beyond the S coding region. These findings underscore the importance of real-time molecular surveillance of SARS-CoV-2 cases for rapidly detecting new emergent viral variants with increased transmissibility and potential to spread globally. Moreover, this study emphasizes the relevance of monitoring the emergence of SARS-CoV-2 recombinant lineages between XBB and BA.2.86 Omicron subvariants and investigating potential determinants of viral transmissibility located outside the S region.

## Data Availability

This study's conclusions derive from examining 6,152 SARS-CoV-2 genomes from Brazil, which have been made publicly accessible via the EpiCov database from GISAID. These genomes were collected after October 1st, 2023, and submissions were recorded up until February 29th, 2024. The data can be accessed at https://doi.org/10.55876/gis8.240321ou. For our investigation of the XDR parental lineages and the phylogeographic analysis, we further included 20,226 global reference sequences collected after September 8th, 2023, and submissions were also recorded up until February 29th, 2024, which are available at https://doi.org/10.55876/gis8.240321ce. The XML files utilized throughout this analysis are openly accessible for review and replication of our methods at https://github.com/larboh-ioc/sars2_xdr.

## ACKNOWLEDGMENTS

We gratefully acknowledge all data contributors, i.e., the originating laboratories responsible for obtaining the specimens and the submitting laboratories for generating the genetic sequence and metadata and sharing via EpiCov database from GISAID, on which this research is based. The authors also wish to thank all the healthcare workers and scientists who have worked hard to deal with this pandemic threat. In addition, we appreciate the support of the Respiratory Viruses Genomic Surveillance Network of the General Laboratory Coordination (CGLab) of the Brazilian Ministry of Health (MoH) and Brazilian Central Laboratory States (LACENs).

## ETHICAL ASPECTS

This study was approved by the Ethics Committee of the FIOCRUZ (CAAE: 68118417.6.0000.5248 and CAAE:32333120.4.0000.5190), which waived the signed informed consent.

## AUTHORS CONTRIBUTIONS

The study was conceived and designed by G.B., F.G.N., and I.A. F.M., R.K., P.C.R., E.D., G.L.W. and F.G.N. contributed to diagnostics and sequencing analysis. K.I. contributed to the instantaneous reproduction number estimations. M.G., F.C.D.C., and W.A.F.D.A. worked on the retrieval and analysis of Brazilian epidemiological data. F.G.N. and M.M.S. contributed to laboratory management and obtaining financial support. The FGSN consortium contributed to the generation of genomic sequences. I.A., K.I., and E.C.P. performed the bioinformatics analysis. I.A. and G.B. wrote the first draft, and all authors contributed and approved the final manuscript.

The representatives of the Fiocruz Genomic Surveillance Network in Brazil are Hazerral Hazerral de Oliveira Santos (LACEN, AL), Ana Flavia Mendonça (LACEN, GO), Gislene Garcia de Castro Lichs (LACEN, MS), Adelino Soares Lima Neto (LACEN, PI), Patricia Brasil (INI, RJ), Andréa Cony (LACEN, RJ), Jayra Juliana Paiva Alves Abrantes (LACEN, RN), Tatiana Schäffer Gregianini (LACEN, RS), Darcita Buerger Rovaris (LACEN, SC), Cliomar Alves dos Santos (LACEN, SE), Franciano Dias Pereira Cardoso (LACEN, TO), Zoraida del Carmen Fernandez Grillo (FIOCRUZ, MS), Adriano Abbud (Instituto Adolfo Lutz, SP), Luana Barbagelata (Instituto Evandro Chagas, PA), Leandro Cavalcante Santos (LACEN, AC), Márcia Socorro Pereira Cavalcante (LACEN, AP), Valnete Andrade (LACEN, PA), Thiago Franco de Oliveira Carneiro (LACEN, PB), Shirlene Telmos Silva de Lima (LACEN, CE), Jórdan Barros da Silva (LACEN, DF), Rodrigo Ribeiro Rodrigues (LACEN, ES), Cliomar Alves dos Santos (LACEN, SE).

## REFERENCES

Altschul, S. F., Gish, W., Miller, W., Myers, E. W., & Lipman, D. J. (1990). Basic local alignment search tool. Journal of Molecular Biology, 215(3), 403–410. 10.1016/s0022-2836(05)80360-2

Anisimova, M., & Gascuel, O. (2006). Approximate likelihood-ratio test for branches: A fast, accurate, and powerful alternative. Systematic Biology, 55(4), 539–552. 10.1080/10635150600755453

Arantes, I., Bello, G., Nascimento, V., Souza, V., da Silva, A., Silva, D., Nascimento, F., Mejía, M., Brandão, M. J., Gonçalves, L., Silva, G., da Costa, C. F., Abdalla, L., Santos, J. H., Ramos, T. C. A., Piantham, C., Ito, K., Siqueira, M. M., Resende, P. C., … Naveca, F. G. (2023). Comparative epidemic expansion of SARS-CoV-2 variants Delta and Omicron in the Brazilian State of Amazonas. Nature Communications, 14(1). 10.1038/s41467-023-37541-6

Arantes, I., Gomes, M., Ito, K., Sarafim, S., Gräf, T., Miyajima, F., Khouri, R., de Carvalho, F. C., de Almeida, W. A. F., Siqueira, M. M., Resende, P. C., Naveca, F. G., & Bello, G. (2024). Spatiotemporal dynamics and epidemiological impact of SARS-CoV-2 XBB lineage dissemination in Brazil in 2023. Microbiology Spectrum, 12(3). 10.1128/spectrum.03831-23

Bouckaert, R., Vaughan, T. G., Barido-Sottani, J., Duchêne, S., Fourment, M., Gavryushkina, A., Heled, J., Jones, G., Kühnert, D., De Maio, N., Matschiner, M., Mendes, F. K., Müller, N. F., Ogilvie, H. A., du Plessis, L., Popinga, A., Rambaut, A., Rasmussen, D., Siveroni, I., … Drummond, A. J. (2019). BEAST 2.5: An advanced software platform for Bayesian evolutionary analysis. PLOS Computational Biology, 15(4), e1006650. 10.1371/journal.pcbi.1006650

Chen, C., Nadeau, S., Yared, M., Voinov, P., Xie, N., Roemer, C., & Stadler, T. (2021). CoV-Spectrum: Analysis of globally shared SARS-CoV-2 data to identify and characterize new variants. Bioinformatics, 38(6), 1735–1737. 10.1093/bioinformatics/btab856

Dezordi, F. Z., Neto, A. M. da S., Campos, T. de L., Jeronimo, P. M. C., Aksenen, C. F., Almeida, S. P., & Wallau, G. L. (2022). ViralFlow: A versatile automated workflow for sars-cov-2 genome assembly, lineage assignment, mutations and intrahost variant detection. Viruses, 14(2), 217. 10.3390/v14020217

Drummond, A. J. (2005). Bayesian coalescent inference of past population dynamics from molecular sequences. Molecular Biology and Evolution, 22(5), 1185–1192. 10.1093/molbev/msi103

Drummond, A. J., Ho, S. Y. W., Phillips, M. J., & Rambaut, A. (2006). Relaxed phylogenetics and dating with confidence. PLoS Biology, 4(5), e88. 10.1371/journal.pbio.0040088

Duchene, S., Featherstone, L., Haritopoulou-Sinanidou, M., Rambaut, A., Lemey, P., & Baele, G. (2020). Temporal signal and the phylodynamic threshold of SARS-CoV-2. Virus Evolution, 6(2). 10.1093/ve/veaa061

Ghafari, M., du Plessis, L., Raghwani, J., Bhatt, S., Xu, B., Pybus, O. G., & Katzourakis, A. (2022). Purifying Selection Determines the Short-Term Time Dependency of Evolutionary Rates in SARS-CoV-2 and pH1N1 Influenza. Molecular Biology and Evolution, 39(2). 10.1093/molbev/msac009

Gouy, M., Guindon, S., & Gascuel, O. (2009). SeaView version 4: A multiplatform graphical user interface for sequence alignment and phylogenetic tree building. Molecular Biology and Evolution, 27(2), 221–224. 10.1093/molbev/msp259

Held, L., & Sabanés Bové, D. (2020). Likelihood and bayesian inference. Springer Berlin Heidelberg. 10.1007/978-3-662-60792-3

Hu, Y., Zou, J., Kurhade, C., Deng, X., Chang, H. C., Kim, D. K., Shi, P.-Y., Ren, P., & Xie, X. (2023). Less neutralization evasion of SARS-CoV-2 BA.2.86 than XBB sublineages and CH.1.1. Emerging Microbes &amp; Infections, 12(2). 10.1080/22221751.2023.2271089

Ito, K., Piantham, C., & Nishiura, H. (2021). Predicted dominance of variant Delta of SARS-CoV-2 before Tokyo Olympic Games, Japan, July 2021. Eurosurveillance, 26(27). 10.2807/1560-7917.es.2021.26.27.2100570

Kaku, Y., Okumura, K., Padilla-Blanco, M., Kosugi, Y., Uriu, K., Hinay, A. A., Jr, Chen, L., Plianchaisuk, A., Kobiyama, K., Ishii, K. J., Zahradnik, J., Ito, J., & Sato, K. (2024). Virological characteristics of the SARS-CoV-2 JN.1 variant. The Lancet Infectious Diseases, 24(2), e82. 10.1016/s1473-3099(23)00813-7

Kalyaanamoorthy, S., Minh, B. Q., Wong, T. K. F., von Haeseler, A., & Jermiin, L. S. (2017). ModelFinder: Fast model selection for accurate phylogenetic estimates. Nature Methods, 14(6), 587–589. 10.1038/nmeth.4285

Katoh, K., Rozewicki, J., & Yamada, K. D. (2017). MAFFT online service: Multiple sequence alignment, interactive sequence choice and visualization. Briefings in Bioinformatics, 20(4), 1160–1166. 10.1093/bib/bbx108

Khan, K., Lustig, G., Römer, C., Reedoy, K., Jule, Z., Karim, F., Ganga, Y., Bernstein, M., Baig, Z., Jackson, L., Mahlangu, B., Mnguni, A., Nzimande, A., Stock, N., Kekana, D., Ntozini, B., van Deventer, C., Marshall, T., Manickchund, N., … Sigal, A. (2023). Evolution and neutralization escape of the SARS-CoV-2 BA.2.86 subvariant. Nature Communications, 14(1). 10.1038/s41467-023-43703-3

Lasrado, N., Collier, A. Y., Hachmann, N. P., Miller, J., Rowe, M., Schonberg, E. D., Rodrigues, S. L., LaPiana, A., Patio, R. C., Anand, T., Fisher, J., Mazurek, C. R., Guan, R., Wagh, K., Theiler, J., Korber, B. T., & Barouch, D. H. (2023). Neutralization escape by SARS-CoV-2 Omicron subvariant BA.2.86. Vaccine, 41(47), 6904–6909. 10.1016/j.vaccine.2023.10.051

Lemey, P., Rambaut, A., Drummond, A. J., & Suchard, M. A. (2009). Bayesian phylogeography finds its roots. PLoS Computational Biology, 5(9), e1000520. 10.1371/journal.pcbi.1000520

Looi, M.-K. (2023). Covid-19: WHO adds JN.1 as new variant of interest. BMJ, 2975. 10.1136/bmj.p2975

Minh, B. Q., Schmidt, H. A., Chernomor, O., Schrempf, D., Woodhams, M. D., von Haeseler, A., & Lanfear, R. (2020). IQ-TREE 2: New models and efficient methods for phylogenetic inference in the genomic era. Molecular Biology and Evolution, 37(5), 1530–1534. 10.1093/molbev/msaa015

Naveca, F. G., Nascimento, V., de Souza, V. C., Corado, A. de L., Nascimento, F., Silva, G., Costa, Á., Duarte, D., Pessoa, K., Mejía, M., Brandão, M. J., Jesus, M., Gonçalves, L., da Costa, C. F., Sampaio, V., Barros, D., Silva, M., Mattos, T., Pontes, G., … Bello, G. (2021). COVID-19 in Amazonas, Brazil, was driven by the persistence of endemic lineages and P.1 emergence. Nature Medicine, 27(7), 1230–1238. 10.1038/s41591-021-01378-7

O’Brien, J. D., Minin, V. N., & Suchard, M. A. (2009). Learning to count: Robust estimates for labeled distances between molecular sequences. Molecular Biology and Evolution, 26(4), 801–814. 10.1093/molbev/msp003

Okada, P., Buathong, R., Phuygun, S., Thanadachakul, T., Parnmen, S., Wongboot, W., Waicharoen, S., Wacharapluesadee, S., Uttayamakul, S., Vachiraphan, A., Chittaganpitch, M., Mekha, N., Janejai, N., Iamsirithaworn, S., Lee, R. T., & Maurer-Stroh, S. (2020). Early transmission patterns of coronavirus disease 2019 (COVID-19) in travellers from Wuhan to Thailand, January 2020. Eurosurveillance, 25(8). 10.2807/1560-7917.es.2020.25.8.2000097

O’Toole, Á., Scher, E., Underwood, A., Jackson, B., Hill, V., McCrone, J. T., Colquhoun, R., Ruis, C., Abu-Dahab, K., Taylor, B., Yeats, C., du Plessis, L., Maloney, D., Medd, N., Attwood, S. W., Aanensen, D. M., Holmes, E. C., Pybus, O. G., & Rambaut, A. (2021). Assignment of epidemiological lineages in an emerging pandemic using the pangolin tool. Virus Evolution, 7(2). 10.1093/ve/veab064

Park, S. W., Sun, K., Abbott, S., Sender, R., Bar-on, Y. M., Weitz, J. S., Funk, S., Grenfell, B. T., Backer, J. A., Wallinga, J., Viboud, C., & Dushoff, J. (2023). Inferring the differences in incubation-period and generation-interval distributions of the Delta and Omicron variants of SARS-CoV-2. Proceedings of the National Academy of Sciences, 120(22). 10.1073/pnas.2221887120

Planas, D., Staropoli, I., Michel, V., Lemoine, F., Donati, F., Prot, M., Porrot, F., Guivel-Benhassine, F., Jeyarajah, B., Brisebarre, A., Dehan, O., Avon, L., Bolland, W. H., Hubert, M., Buchrieser, J., Vanhoucke, T., Rosenbaum, P., Veyer, D., Péré, H., … Schwartz, O. (2024). Distinct evolution of SARS-CoV-2 Omicron XBB and BA.2.86/JN.1 lineages combining increased fitness and antibody evasion. Nature Communications, 15(1). 10.1038/s41467-024-46490-7

Qu, P., Xu, K., Faraone, J. N., Goodarzi, N., Zheng, Y.-M., Carlin, C., Bednash, J. S., Horowitz, J. C., Mallampalli, R. K., Saif, L. J., Oltz, E. M., Jones, D., Gumina, R. J., & Liu, S.-L. (2024). Immune evasion, infectivity, and fusogenicity of SARS-CoV-2 BA.2.86 and FLip variants. Cell, 187(3), 585-595.e6. 10.1016/j.cell.2023.12.026

R Core Team (2020) R: A Language and Environment for Statistical Computing. R Foundation for Statistical Computing,Vienna, Austria. https://www.r-project.org/

Rambaut, A. (2000). Estimating the rate of molecular evolution: Incorporating non-contemporaneous sequences into maximum likelihood phylogenies. Bioinformatics, 16(4), 395–399. 10.1093/bioinformatics/16.4.395

Rambaut, A., Drummond, A. J., Xie, D., Baele, G., & Suchard, M. A. (2018). Posterior summarization in bayesian phylogenetics using tracer 1.7. Systematic Biology, 67(5), 901–904. 10.1093/sysbio/syy032

Rambaut, A., Holmes, E. C., O’Toole, Á., Hill, V., McCrone, J. T., Ruis, C., du Plessis, L., & Pybus, O. G. (2020). A dynamic nomenclature proposal for SARS-CoV-2 lineages to assist genomic epidemiology. Nature Microbiology, 5(11), 1403–1407. 10.1038/s41564-020-0770-5

Rambaut, A., Lam, T. T., Max Carvalho, L., & Pybus, O. G. (2016). Exploring the temporal structure of heterochronous sequences using TempEst (formerly Path-O-Gen). Virus Evolution, 2(1), vew007. 10.1093/ve/vew007

Resende, P. C., Delatorre, E., Gräf, T., Mir, D., Motta, F. C., Appolinario, L. R., Paixão, A. C. D. da, Mendonça, A. C. da F., Ogrzewalska, M., Caetano, B., Wallau, G. L., Docena, C., Santos, M. C. dos, de Almeida Ferreira, J., Sousa Junior, E. C., Silva, S. P. da, Fernandes, S. B., Vianna, L. A., Souza, L. da C., … Siqueira, M. M. (2021). Evolutionary dynamics and dissemination pattern of the sars-cov-2 lineage B.1.1.33 during the early pandemic phase in Brazil. Frontiers in Microbiology, 11. 10.3389/fmicb.2020.615280

Stadler, T., Kühnert, D., Bonhoeffer, S., & Drummond, A. J. (2012). Birth–death skyline plot reveals temporal changes of epidemic spread in HIV and hepatitis C virus (HCV). Proceedings of the National Academy of Sciences, 110(1), 228–233. 10.1073/pnas.1207965110

Suchard, M. A., Lemey, P., Baele, G., Ayres, D. L., Drummond, A. J., & Rambaut, A. (2018). Bayesian phylogenetic and phylodynamic data integration using BEAST 1.10. Virus Evolution, 4(1). 10.1093/ve/vey016

Tamura, T., Ito, J., Uriu, K., Zahradnik, J., Kida, I., Anraku, Y., Nasser, H., Shofa, M., Oda, Y., Lytras, S., Nao, N., Itakura, Y., Deguchi, S., Suzuki, R., Wang, L., Begum, M. M., Kita, S., Yajima, H., Sasaki, J., … Sato, K. (2023). Virological characteristics of the SARS-CoV-2 XBB variant derived from recombination of two Omicron subvariants. Nature Communications, 14(1). 10.1038/s41467-023-38435-3

Tamura, T., Mizuma, K., Nasser, H., Deguchi, S., Padilla-Blanco, M., Oda, Y., Uriu, K., Tolentino, J. E. M., Tsujino, S., Suzuki, R., Kojima, I., Nao, N., Shimizu, R., Wang, L., Tsuda, M., Jonathan, M., Kosugi, Y., Guo, Z., Hinay, A. A., Jr., … Sato, K. (2024). Virological characteristics of the SARS-CoV-2 BA.2.86 variant. Cell Host &amp; Microbe, 32(2), 170-180.e12. 10.1016/j.chom.2024.01.001

Tay, J. H., Porter, A. F., Wirth, W., & Duchene, S. (2022). The emergence of sars-cov-2 variants of concern is driven by acceleration of the substitution rate. Molecular Biology and Evolution, 39(2). 10.1093/molbev/msac013

Turakhia, Y., Thornlow, B., Hinrichs, A., McBroome, J., Ayala, N., Ye, C., Smith, K., De Maio, N., Haussler, D., Lanfear, R., & Corbett-Detig, R. (2022). Pandemic-scale phylogenomics reveals the SARS-CoV-2 recombination landscape. Nature, 609(7929), 994–997. 10.1038/s41586-022-05189-9

Uriu, K., Ito, J., Kosugi, Y., Tanaka, Y. L., Mugita, Y., Guo, Z., Hinay, A. A., Jr, Putri, O., Kim, Y., Shimizu, R., Begum, M. M., Jonathan, M., Saito, A., Ikeda, T., & Sato, K. (2023). Transmissibility, infectivity, and immune evasion of the SARS-CoV-2 BA.2.86 variant. The Lancet Infectious Diseases, 23(11), e460–e461. 10.1016/s1473-3099(23)00575-3

Wang, L.-G., Lam, T. T.-Y., Xu, S., Dai, Z., Zhou, L., Feng, T., Guo, P., Dunn, C. W., Jones, B. R., Bradley, T., Zhu, H., Guan, Y., Jiang, Y., & Yu, G. (2019). Treeio: An R package for phylogenetic tree input and output with richly annotated and associated data. Molecular Biology and Evolution, 37(2), 599–603. 10.1093/molbev/msz240

Wang, Q., Guo, Y., Liu, L., Schwanz, L. T., Li, Z., Nair, M. S., Ho, J., Zhang, R. M., Iketani, S., Yu, J., Huang, Y., Qu, Y., Valdez, R., Lauring, A. S., Huang, Y., Gordon, A., Wang, H. H., Liu, L., & Ho, D. D. (2023). Antigenicity and receptor affinity of SARS-CoV-2 BA.2.86 spike. Nature, 624(7992), 639–644. 10.1038/s41586-023-06750-w

Wang, X., Lu, L., & Jiang, S. (2024). SARS-CoV-2 evolution from the BA.2.86 to JN.1 variants: Unexpected consequences. Trends in Immunology, 45(2), 81–84. 10.1016/j.it.2024.01.003

WHO. (2020). Tracking SARS-CoV-2 variants. Tracking SARS-CoV-2 Variants. https://www.who.int/activities/tracking-SARS-CoV-2-variants

WHO. (2023, December 13). Statement on the antigen composition of COVID-19 vaccines. World Health Organization. https://www.who.int/news/item/13-12-2023-statement-on-the-antigen-composition-of-covid-19-vaccines

Wickham, H. (2016). ggplot2. Springer International Publishing. 10.1007/978-3-319-24277-4

Yang, S., Yu, Y., Xu, Y., Jian, F., Song, W., Yisimayi, A., Wang, P., Wang, J., Liu, J., Yu, L., Niu, X., Wang, J., Wang, Y., Shao, F., Jin, R., Wang, Y., & Cao, Y. (2024). Fast evolution of SARS-CoV-2 BA.2.86 to JN.1 under heavy immune pressure. The Lancet Infectious Diseases, 24(2), e70–e72. 10.1016/s1473-3099(23)00744-2

Yu, G. (2020). Using ggtree to Visualize Data on Tree-Like Structures. Current Protocols in Bioinformatics, 69(1). 10.1002/cpbi.96

Zhang, L., Kempf, A., Nehlmeier, I., Cossmann, A., Richter, A., Bdeir, N., Graichen, L., Moldenhauer, A.-S., Dopfer-Jablonka, A., Stankov, M. V., Simon-Loriere, E., Schulz, S. R., Jäck, H.-M., Čičin-Šain, L., Behrens, G. M. N., Drosten, C., Hoffmann, M., & Pöhlmann, S. (2024). SARS-CoV-2 BA.2.86 enters lung cells and evades neutralizing antibodies with high efficiency. Cell, 187(3), 596-608.e17. 10.1016/j.cell.2023.12.025

