## Supplementary_Material for "Rapid spread of the SARS-CoV-2 Omicron XDR lineage derived from recombination between XBB and BA.2.86 subvariants circulating in Brazil in late 2023"

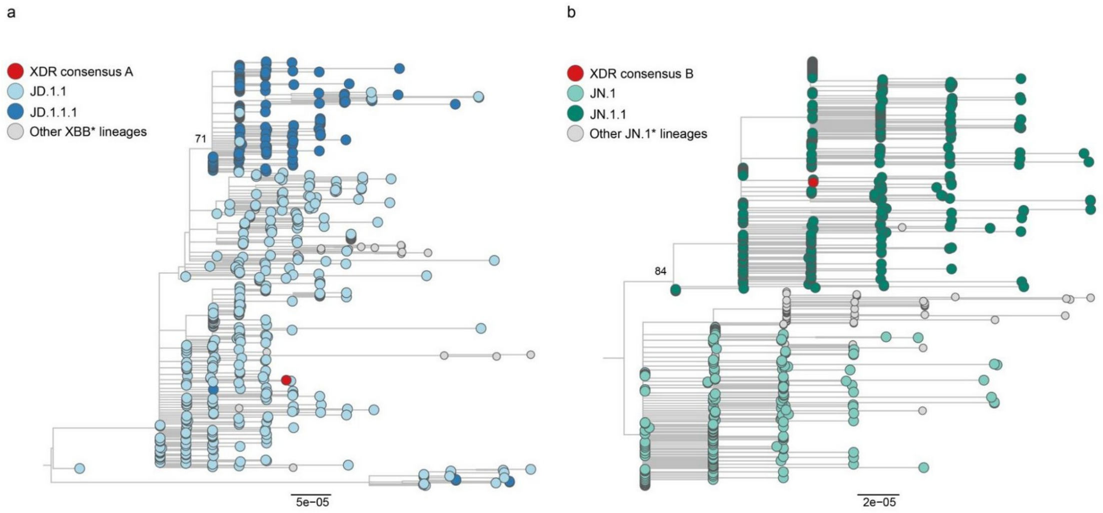


**Supplementary Figure 1: Identification of XDR parental lineages**. The figure depicts the ML trees inferred with a subgenomic consensus XDR sequence (A: positions 1 to 11,043; B: positions 11,726 to 29,421) and its most closely associated XBB (**a**) and BA.2.86 (**b**) subvariant genome sequences. The statistical support (aLRT) of key nodes in the trees are annotated. The trees are rooted at their midpoints and drawn according to the scale at the bottom of each panel.

**Supplementary Table 1: Evolution of the XDR $R_e$ in Brazil**

| Time Interval | $R_e$ (Median) | $R_e$ (95% HPD) |
|---|---|---|
| 2023-11-08 to 2023-12-01 | 1.52 | 1.08 - 2.19 |
| 2023-12-02 to 2023-12-25 | 1.42 | 1.16 - 2.35 |
| 2023-12-26 to 2024-01-17 | 1.02 | 0.80 - 1.29 |
| 2024-01-18 to 2024-02-10 | 0.86 | 0.42 -0.99 |

The table details the Bayesian genomic estimations of the medium value and 95% HPD interval of the effective reproductive number ($R_e$) of the XDR lineage circulating in Brazil between November 2023 and February 2024.
